# Diagnostic performance of the fragmented QRS complex on electrocardiogram for detecting myocardial scars assessed by 3.0 Tesla cardiac magnetic resonance imaging

**DOI:** 10.1101/2024.03.23.24304647

**Authors:** Kasin Viriyanukulvong, Nonthikorn Theerasuwipakorn, Wanwarang Wongcharoen, Paisit Kosum, Ronpichai Chokesuwattanaskul

## Abstract

**Background:** Fragmented QRS complex (f-QRS) on a 12-lead electrocardiogram (EKG) with a 0.15-100 or 150 Hz low-pass filter is known to be related to ischemic myocardial scars. Cardiac magnetic resonance (CMR) imaging enhances tissue characterization capability resulting in a better myocardial scar assessment over other noninvasive imaging modalities. However, the diagnostic values of f-QRS on non-ischemic scars and f-QRS from EKG with a 015-40 Hz low-pass filter (routine filter in clinical practice) are unknown. This study aims to evaluate the diagnostic performance of f-QRS (from EKG with 0.15-40 and 0.15-100 Hz low-pass filters) for detecting any myocardial scars (both ischemic and non-ischemic) assessed by 3.0 Tesla CMR.

**Methods:** This cross-sectional study included patients who underwent a 3.0 Tesla CMR scan from May 2020 to May 2023. A 12-lead EKG with 0.15-40 and 0.15-100 Hz low-pass filters, performed on the same day of the CMR scan, was assessed for the presence of f-QRS. The ECG leads were divided into 3 categories (e.g., anterior leads V1-V4; lateral leads I, aVL, V5-V6; and inferior leads II, III, aVF). The f-QRS was defined as the presence of R’ wave or notching in the nadir of the S wave in 2 contiguous leads. The primary outcome was the diagnostic performance of f-QRS from EKG in myocardial scar detection in the corresponding left ventricle (LV) segments. The secondary outcomes were to compare the diagnostic performance of f-QRS in detecting ischemic scars and non-ischemic scars, the diagnostic performance between f-QRS diagnosed from 0.15-40 and 0.15-100 Hz low-pass filters, and the diagnostic performance of f-QRS presented in 2 consecutive leads and f-QRS presented in solitary lead.

**Results:** The study involved 1,692 participants with a median age of 67 (IQR: 59-85) years old and 52.5% males. Myocardial scars were found in 826 (49%) participants. Male, history of CAD, and myocardial scars were significantly more frequent in the participants with f- QRS (59.4% vs 46.0%, 26.4% vs 20.6%, and 48.9% vs 37.3%, respectively), while median LVEF was lower (61%, IQR 47, 66 vs 62%, IQR 55, 68; p < 0.001). The sensitivity, specificity, positive predictive value, negative predictive value, and AUC of f-QRS from EKG with 0.15-100 Hz low-pass filter for detecting myocardial scars were 25.6%, 88.7%, 45.1%, 76.8%, and 0.57 for anterior segments; 22.1%, 91.5%, 36.8%, 84.1%, and 0.57 for lateral segments; and 42.9%, 63.4%, 36.9, 68.9%, and 0.53 for inferior segments. The sensitivity, PPV, and positive likelihood ratio (LR+) of f-QRS were higher for detecting non-ischemic scars while specificity, NPV, negative likelihood ratio (LR-), and AUC were not significantly different. The f-QRS from 0.15-100 Hz showed a higher sensitivity but lower specificity, PPV, and LR+ for all LV segments. The f-QRS presented in the solitary lead showed a higher sensitivity with a lower specificity, PPV, and LR+.

**Conclusion:** This study demonstrates a high specificity and negative predictive value of f- QRS from a 12-lead EKG with 0.15-40 and 0.15-100 Hz low-pass filters in diagnosing myocardial scars when correlated to the corresponding LV segments.

**Graphical Abstract:** 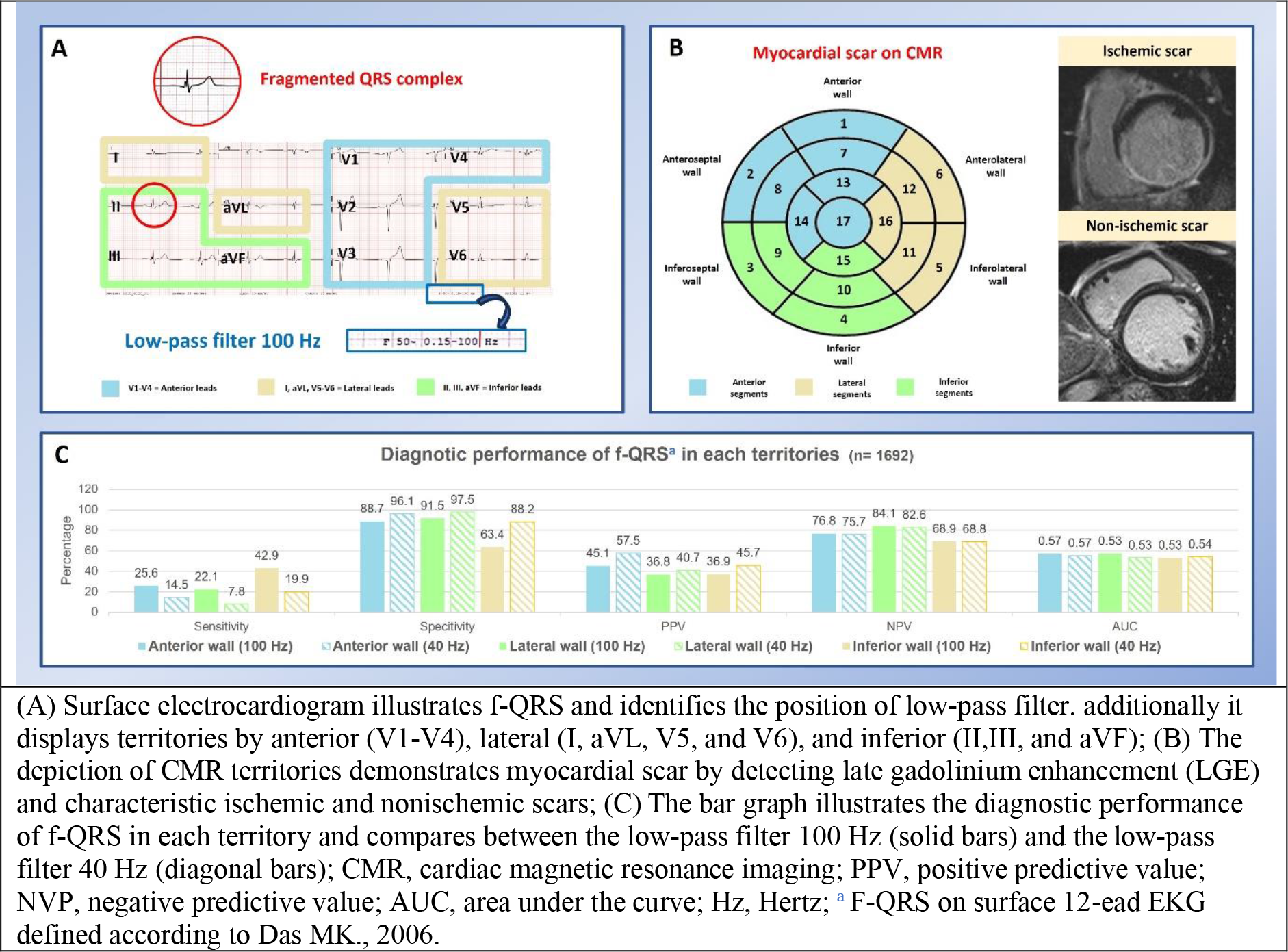

## INTRODUCTION

The 12-lead electrocardiogram (EKG) is a fundamental tool in diagnosing various cardiac conditions and is universally available. (1) Abnormal electrical conduction through areas of fibrosing myocardium (e.g., myocardial scars) can lead to the expression of the fragmented QRS complex (f-QRS) on the EKG waveform. (2, 3) In 2006, Das and colleagues established a correlation between f-QRS and myocardial scars detected by single photon emission computed tomography (SPECT) in patients with coronary artery disease (CAD).

Their study demonstrated higher sensitivity and negative predictive value (NPV) of f-QRS compared to Q waves. (4) Moreover, the presence of f-QRS correlated with adverse clinical outcomes such as mortality, CAD, and heart failure. (5, 6) However, previous publications primarily focused on the scars assessed by nuclear imaging, limiting data from cardiac magnetic resonance (CMR) imaging which is considered the gold standard noninvasive imaging modality for assessing myocardial scars. (7) Furthermore, the diagnostic performance of f-QRS in detecting myocardial scars other than ischemic scars was still unknown. To enhance f-QRS detection sensitivity, previous studies typically set a low-pass filter between 0.15-100 and 0.15-150 Hz which was not routinely used in clinical practice. (8)

Therefore, we aim to assess the diagnostic performance of f-QRS for detecting any myocardial scars (both ischemic and non-ischemic) assessed by CMR. In addition, this study also compares the performance of f-QRS from each filter range (0.15-40 and 0.15-100 Hz low-pass filters) and 2 consecutive leads vs solitary lead.

## MATERIALS AND METHODS

This cross-sectional study enrolled patients aged 18 years or older who underwent a 3.0 Tesla CMR scan with perfusion protocol from May 2020 to May 2023 at King Chulalongkorn Memorial Hospital, Bangkok, Thailand. The key exclusion criteria were patients with (i) a recent myocardial infarction within 30 days, (ii) pre-excitation syndrome, (iii) uninterpretable EKG, and (iv) inadequate CMR image quality for scar assessment. The baseline characteristics of the participants were retrieved from medical electronic databases. The study protocol was reviewed and approved by the Institutional Research Committee, Faculty of Medicine, Chulalongkorn University (IRB No. 0660/66).

### Electrocardiogram collection and interpretation

A surface 12-lead EKG with 0.15-40 and 0.15-100 Hz low-pass filters was performed on the same day of the CMR scan using a digital machine, the PageWriter TC 70 (Philips, Eindhoven, the Netherlands). The EKG was run with a paper speed of 25 mm/s and an amplitude of 10 mm/mV. The EKG waveforms were electrically transmitted to the Tracemaster system and analyzed on automatic analysis software (IntelliSpace ECG, Philips, Eindhoven, The Netherlands) with manual correction. EKG with each filter range was assessed independently for f-QRS by two cardiologists (K.V. and R.C.) blinded to the CMR results. An electrophysiologist (W.W.) adjudicated the EKG findings in case of discordancy. The f-QRS is defined as the presence of an additional R wave (R’) or notching in the nadir of the S wave in 2 contiguous leads (4, 9). The f-QRS in a solitary lead was also gathered owing to the discrete nature of non-ischemic cardiomyopathy scars, which may not align with coronary artery territories and might have the potential to detect smaller scar areas in non-ischemic conditions.

### CMR image acquisition and post-processing

All enrolled patients were scanned with perfusion protocol using a 3.0 Tesla scanner (Magnetom Vida, Siemens Healthineers, Erlangen, Germany) with an 18-channel cardiac phased array receiver. Gadobutrol (Gadovist, Bayer Healthcare, Leverkusen, Germany) or Gadoterate meglumine (Dotarem; Guerbet, Villepinte, France) with a total dose of 0.15 mmol/kg was injected during first-pass perfusion images acquisition. Late gadolinium enhancement (LGE) images with optimal inversion time (Ti) received from Ti scout were acquired 5-10 minutes after gadolinium injection. Spoiled gradient echo with phase-sensitive reconstruction (PSIR) acquired during breath holding in 2-chamber, 3-chamber, 4-chamber, and short-axis stack covering the whole ventricle was used as a primary pulse sequence for LGE images acquisition and interpretation. If patients could not perform breath holding or significant arrhythmia occurred, single-shot, non-breath holding, steady-state free precession pulse sequence was utilized for LGE images instead. Each slice was configured with a 6 mm thickness without a slice gap. Myocardial scar was diagnosed when LGE was visualized in two orthogonal planes and further classified into ischemic scars (subendocardial and transmural) and non-ischemic scars (mid-wall, subepicardial, and patchy). (10) Post-processing software (Syngo.via, Siemens Healthineers, Erlangen, Germany) was used to analyze images. Data regarding the presence of myocardial scars was retrieved from the CMR reports validated by experienced CMR imaging specialists.

### EKG leads and LV segmentation

EKG leads and LV segments were divided into 3 groups correlated to each other.

EKG leads V1-V4 were categorized as anterior leads and interpreted as corresponding to the myocardial scar in anterior segments of LV (basal anterior, basal anteroseptal, mid anterior, mid anteroseptal, apical anterior, apical septal, and apical cap segments). EKG leads I, aVL, and V5-V6 were categorized as lateral leads and interpreted as corresponding to the myocardial scar in lateral segments of LV (basal anterolateral, basal inferolateral, mid anterolateral, mid inferolateral, and apical lateral segments). EKG leads II, III, and aVF were categorized as inferior leads and interpreted as corresponding to the myocardial scar in inferior segments of LV (basal inferoseptal, basal inferior, mid inferoseptal, mid inferior, and apical inferior segments). **[Graphical Abstract]**

### Primary and secondary outcomes

The primary outcome was the diagnostic performance of f-QRS from EKG with 0.15- 100 Hz low-pass filter in myocardial scar detection in the corresponding left ventricle (LV) segments which was described by sensitivity, specificity, positive predictive value (PPV), NPV, positive (LR+) and negative likelihood ratio (LR-), and area under the receiver operating characteristic (ROC) curve (AUC). The secondary outcomes were (i) to compare the diagnostic performance of f-QRS in detecting ischemic scars and non-ischemic scars, (ii) to compare the diagnostic performance between f-QRS diagnosed from 0.15-40 (routine) and 0.15-100 Hz (standard) low-pass filters, and (iii) to compare the diagnostic performance of f- QRS presented in 2 consecutive leads and f-QRS presented in solitary lead.

### Statistical analysis

Categorical data was presented as frequency with percentage and analyzed with the Chi-square or Fisher’s exact tests as appropriate. Continuous data was displayed as mean with standard deviation (SD) or median with interquartile range (IQR) and analyzed with independent t-test or Mann-Whitney U test as appropriate. The sensitivity, specificity, PPV, NPV, LR+, and LR- were calculated to demonstrate the diagnostic performance of f-QRS. ROC curve was employed to investigate the relationship between f-QRS and myocardial scar which presented as AUC. Two assessors of EKG were tested for inter- and intra-observer reliability. A p-value of less than 0.05 is considered statistically significant. The statistical analyses were conducted using the IBM SPSS software, version 29 for Windows.

## RESULTS

### Study population and baseline characteristics

A total of 1,728 patients were identified by searching the electrical database from May 2020 to May 2023. Twenty-four patients with recent-onset myocardial infarction, 6 patients with uninterpretable EKG, and 6 patients with inadequate CMR image quality were excluded, leaving 1,692 participants for the analyses **[Figure 1]**. The median age was 67 (IQR: 59, 85) years old and 52.5% were male. 826 (49%) participants exhibited f-QRS on EKG. Hypertension, diabetes mellitus, and dyslipidemia were present in 1,082 (64%), 517 (30.6%), and 1,107 (65.5%) participants, respectively. The median estimated glomerular filtration rate (eGFR) was 78 (IQR: 61, 91) mL/min/1.73m². A history of CAD was found in 396 (23.4%) participants with 13.3% and 2.2% who had experienced percutaneous coronary intervention and coronary artery bypass graft surgery, respectively. The median left ventricular ejection fraction (LVEF) was 61% (IQR; 52, 67). 727 (43%) participants revealed myocardial scars on CMR imaging (ischemic scars 25.7% and non-ischemic scars 19.4%).

**Figure 1.**
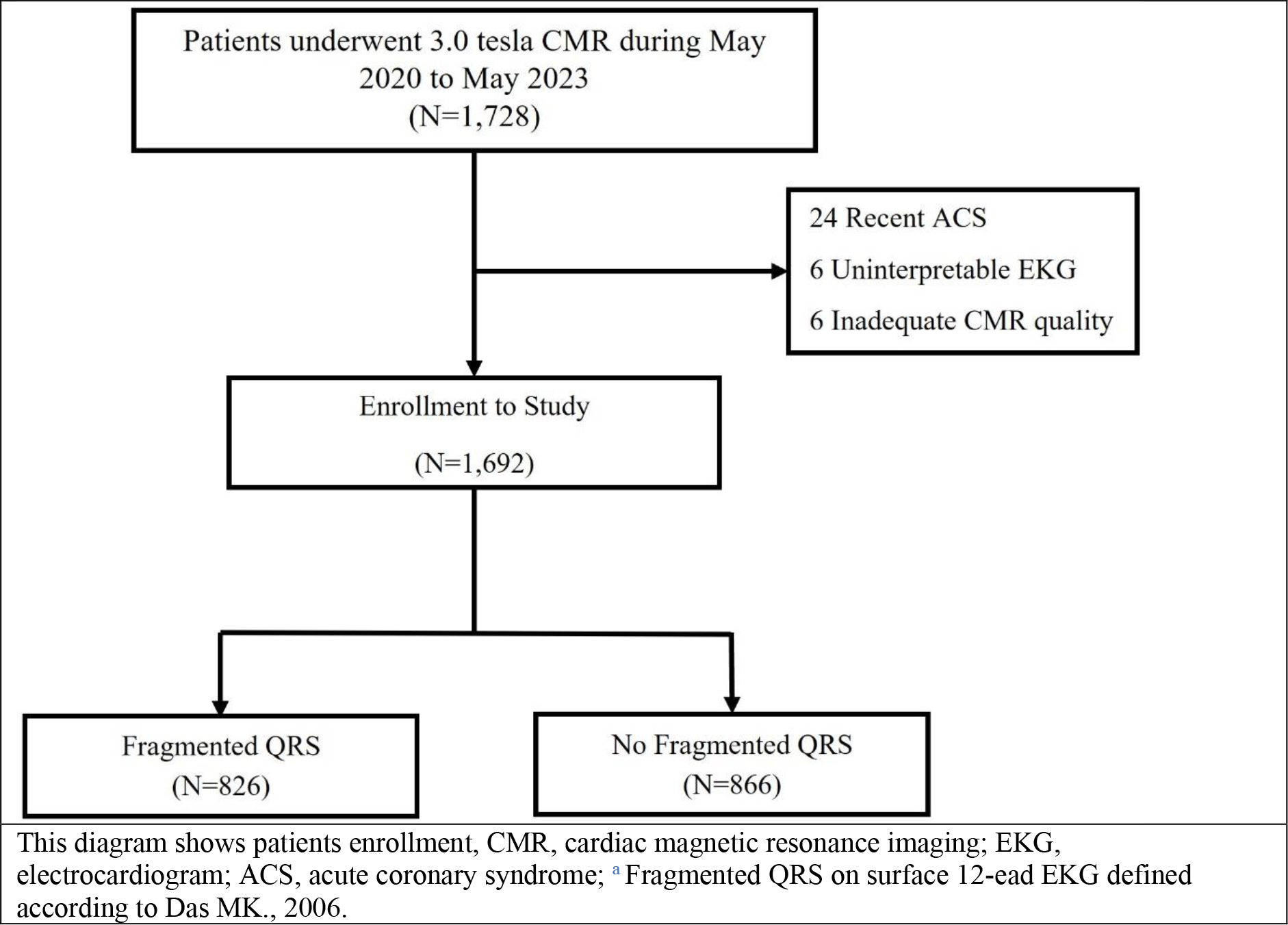
**Study profile**

Myocardial scars were present in the anterior segments in 449 (26.5%) participants, the lateral segments in 308 (18.2%) participants, and the inferior segments in 564 (33.3%) participants. **[Table 1]**

**Table 1.** Baseline characteristics.

|  | Total<br>(N=1692) | No f-QRS<br>(N=866) | f-QRS<br>(N=826) | p-value |
| --- | --- | --- | --- | --- |
| Male, n (%) | 889 (52.5) | 389 (46) | 491 (59.4) | <0.001 |
| Age (years), median (IQR) | 67 (59-85) | 68 (60-75) | 67 (59-75) | 0.62 |
| Hypertension, n (%) | 1082 (64) | 563 (65) | 519 (62.8) | 0.35 |
| Diabetes mellitus, n (%) | 517 (30.6) | 264 (30.5) | 253 (30.6) | 0.95 |
| Dyslipidemia, n (%) | 1107 (65.5) | 572 (66.1) | 535 (64.8) | 0.56 |
| eGFR (mL/min/1.73m<sup>2</sup>), median (IQR) | 78 (61-91) | 79 (60.5-92) | 77 (61-90) | 0.12 |
| CKD Stage, n (%) |  |  |  | 0.32 |
| Stage 1 | 471 (27.9) | 257 (29.7) | 214 (25.9) |  |
| Stage 2 | 818 (48.4) | 400 (46.2) | 418 (50.7) |  |
| Stage 3 | 347 (20.5) | 183 (21.2) | 164 (19.9) |  |
| Stage 4 | 28 (1.7) | 13 (1.5) | 15 (1.8) |  |
| Stage 5 | 26 (1.5) | 12 (1.4) | 14 (1.7) |  |
| Coronary artery disease, n (%) | 396 (23.4) | 178 (20.6) | 218 (26.4) | 0.005 |
| Previous PCI, n (%) | 225 (13.3) | 107 (12.4) | 118 (14.3) | 0.24 |
| Previous CABG, n (%) | 37 (2.2) | 18 (2.1) | 19 (2.3) | 0.75 |
| LVEF (%), median (IQR) | 61 (52-67) | 62 (55-68) | 61 (47-66) | <0.001 |
| Presence of scar, n (%) | 727 (43) | 323 (37.3) | 404 (48.9) | <0.001 |
| Presence of infarct scar, n (%) | 434 (25.7) | 182 (21.0) | 252 (30.5) | <0.001 |
| Presence of non-infarct scar, n (%) | 329 (19.4) | 150 (17.3) | 179 (21.7) | 0.02 |
| Data are presented as median (interquartile range) of frequency and percentage. eGFR, estimated glomerular filtration rate; CKD, chronic kidney disease; LVEF, left ventricular ejection fraction; PCI, percutaneous coronary intervention; CABG, coronary artery bypass graft; CMR, cardiac magnetic resonance imaging; <sup>a</sup>A scar refer to detected late gadolinium enhancement (LGE) |  |  |  |  |

Male, history of CAD, and myocardial scars were significantly more frequent in the participants with f-QRS (59.4% vs 46%, p-value < 0.001 for male; 26.4% vs 20.6%, p-value 0.005 for CAD; and 48.9% vs 37.3%, p-value < 0.001 for the presence of scars), while median LVEF was lower (61%, IQR 47, 66 vs 62%, IQR 55, 68; p < 0.001). Other baseline characteristics were not significantly different between the 2 groups.

The interobserver reliability for the presence of f-QRS was 0.78 (95% CI: 0.53, 0.94) as determined by Fleiss’Kappa coefficient. The overall intraobserver for the presence of f- QRS was 0.78 (95% CI: 0.61-0.90) using the intraclass correlation coefficient (ICC).

### Primary outcome

The sensitivity, specificity, PPV, NPV, LR+, LR-, and AUC of f-QRS from EKG with 0.15-100 Hz low-pass filter for detecting myocardial scars were 25.6%, 88.7%, 45.1%, 76.8%, 2.27, 0.84, and 0.57 for anterior segments; 22.1%, 91.5%, 36.8%, 84.1%, 2.61, 0.85, and 0.57 for lateral segments; and 42.9%, 63.4%, 36.9, 68.9%, 1.17, 0.9, and 0.53 for inferior segments. **[Table 2 and Graphical Abstract]**

**Table 2.** Primary outcomes.

|  | Sensitivity<br>(95%CI) | Specificity<br>(95%CI) | PPV<br>(95%CI) | NVP<br>(95%CI) | LR+<br>(95%CI) | LR-<br>(95%CI) | AUC<br>(95%CI) |
| --- | --- | --- | --- | --- | --- | --- | --- |
| Anterior leads and LV segments |  |  |  |  |  |  |  |
| 40-Hz filter | 14.5 (11.4-18.1) | 96.1 (94.9-97.1) | 57.5 (47.9-66.8) | 75.7 (73.5-77.8) | 3.75 (1.92-5.58) | 0.89 (0.82-0.96) | 0.55 (0.54-0.57) |
| 100-Hz filter | 25.6 (21.6-29.9) | 88.7 (86.8-90.4) | 45.1 (38.9-51.4) | 76.8 (74.5-78.9) | 2.27 (1.52-3.02) | 0.84 (0.81-0.87) | 0.57 (0.55-0.59) |
| Lateral leads and LV segments |  |  |  |  |  |  |  |
| 40-Hz filter | 7.8 (5.1-11.4) | 97.5 (96.5-98.2) | 40.7 (28.1-54.3) | 82.6 (80.7-84.4) | 3.08 (1.74-4.42) | 0.95 (0.93-0.97) | 0.53 (0.51-0.54) |
| 100-Hz filter | 22.1 (17.6-27.1) | 91.5 (90-93) | 36.8 (29.8-44.1) | 84.1 (82.1-85.9) | 2.61 (1.79-3.43) | 0.85 (0.82-0.88) | 0.57 (0.54-0.59) |
| Inferior leads and LV segments |  |  |  |  |  |  |  |
| 40-Hz filter | 19.9 (16.6-23.4) | 88.2 (86.2-90) | 45.7 (39.4-52.2) | 68.8 (66.3-71.1) | 1.68 (1.24-2.12) | 0.91 (0.87-0.95) | 0.54 (0.52-0.56) |
| 100-Hz filter | 42.9 (38.8-47.1) | 63.4 (60.5-66.2) | 36.9 (33.2-40.8) | 68.9 (66-71.8) | 1.17 (0.96-1.38) | 0.90 (0.85-0.95) | 0.53 (0.51-0.56) |
| Diagnostic performance of f-QRS for detecting myocardial scars; LR, likelihood ratio; PPV, positive predictive value; NVP, negative predictive value; OR, odd ratio; CI, confidence interval; Hz, Hertz. |  |  |  |  |  |  |  |

### Secondary outcomes

#### f-QRS for detecting ischemic and non-ischemic scars

The sensitivity, PPV, and LR+ of f-QRS were higher for detecting non-ischemic scars compared to ischemic scars while specificity, NPV, LR-, and AUC were not significantly different. These findings were not only similar for all EKG categories and LV segments but also independent of low-pass filters. **[Supplementary Table 1]**

#### f-QRS from 0.15-40 (routine) vs 0.15-100 Hz (standard) low-pass filter

The f-QRS from 0.15-100 Hz showed a higher sensitivity but lower specificity, PPV, and LR+ for all EKG categories and LV segments. NPV, LR-, and AUC were not significantly different between the 2 filters. **[Table 2]**

#### f-QRS presented in 2 consecutive leads vs presented in the solitary lead

Compared with standard 2 consecutive leads, f-QRS presented in the solitary lead showed a higher sensitivity with a lower specificity, PPV, and LR+. NPV and AUC were not significantly different. LR- of f-QRS in the solitary lead was lower in anterior and lateral segments but higher in the inferior segment. **[Supplementary table 2]**

## DISCUSSION

This study investigated the diagnostic performance of f-QRS for detecting any myocardial scars (both ischemic and non-ischemic) assessed by CMR. To the best of our knowledge, the current study included the largest size of participants and the only study that assessed myocardial scars by using 3.0 Tesla CMR. The f-QRS from EKG with 0.15-100 Hz low-pass filter was found in half of the total participants. Participants with f-QRS had more underlying CAD, lower LVEF, and more myocardial scars (both ischemic and non-ischemic).

This study demonstrated unsatisfactory diagnostic performance of f-QRS from 0.15- 100 Hz low-pass filter EKG for detecting myocardial scars in every EKG lead and LV segment with a sensitivity of 22-43%, PPV of 36-45%, LR+ of 1.17-2.61, LR- of 0.84-0.9, and AUC of 0.53-0.57. The specificity (63-91%) and NPV (69-84%) were the only 2 parameters that showed desirable values. Surprisingly, this study’s results were different from most of the previous publications that reported a high sensitivity (78-86%), specificity (65- 93%), PPV (39-84%), and NPV (88-91%). (4, 7, 11, 12) All of these studies utilized stress SPECT with Technetium-99m (^99m^Tc) sestamibi as a modality for myocardial scar assessment. In contrast to our study, we used CMR which is considered the gold standard noninvasive test for evaluating myocardial scars. (13, 14) CMR provided better spatial resolution resulting in a higher accuracy for detecting myocardial scars. (15) In addition, SPECT with ^99m^Tc sestamibi was known for its limitation in classifying true myocardial scars and hibernating viable myocardium. Myocardial scars were overestimated from SPECT with ^99m^Tc sestamibi when compared with SPECT with Thallium-201 (^201^TI), positron emission tomography with ^18^F-Fludeoxyglucose, and CMR. (16–18) The limitations of SPECT with ^99m^Tc sestamibi may be the explanation for the different results in our study. One study diagnosing myocardial scars with 1.5 Tesla CMR also reported higher sensitivity (68%) and PPV (95%) with lower specificity (30%) and NPV (5%) compared to our study. (19) However, the previous study included patients with a history of myocardial infarction which was a dissimilar population to our study. Compared to 3.0 Tesla CMR, likewise, 1.5 Tesla

CMR has a lower signal-to-noise ratio and blood-to-myocardial contrast causing a poorer accuracy for myocardial scar identification. Another noteworthy finding in our study is that detecting f-QRS in the inferior territory exhibited higher sensitivity but lower specificity compared to other regions. One hypothesis is that other related factors may influence the presence of f-QRS in interior territories. A study of f-QRS in healthy adults found that they are most commonly detected as inferior leads and associated with left deviation of the frontal QRS axis rather than myocardial scars, (20) Additionally, f-QRS in inferior leads may be associated with epicardial fat and metabolic syndrome. This relationship has been noted in the previous study. (21)

Almost all of the previous publications investigated the diagnostic performance of f- QRS only in the context of ischemic scars (e.g., patients with coronary artery disease and myocardial infarction). Our study explored the performance of f-QRS in both ischemic and non-ischemic scars. This study demonstrated a higher sensitivity, PPV, and LR+ of f-QRS for detecting non-ischemic scars with an indifferent specificity, NPV, LR-, and AUC. These findings were consistent in all EKG categories, LV segments, and low-pass filter ranges.

Even with higher performance, unfortunately, f-QRS was inadequate to use as a non-ischemic scar indicator. Unlike our findings, two studies that examined the value of f-QRS in patients with nonischemic dilated cardiomyopathy and hypertrophic cardiomyopathy found an excellent association between f-QRS and myocardial scars. (22, 23) In contrast to our study which included 329 patients with non-ischemic scars (19% of overall participants), these 2 studies included only 17 and 39 patients in their analysis. Moreover, the population in these studies was different from our study.

According to its higher sensitivity, an EKG with 0.15-100 or 0.15-150 Hz low-pass filters was theoretically suitable and considered a standard filter for detecting f-QRS. (8) However, these filter ranges were not acquired in routine clinical practice which usually used a lower low-pass filter (e.g., 0.15-40 Hz). This was the first study that compared the performance of f-QRS from EKG with 0.15-40 (routine) and 0.15-100 Hz (standard) low-pass filters. The f-QRS from EKG with a standard 0.15-100 Hz low-pass filter expectedly demonstrated a higher sensitivity with lower specificity, PPV, and LR+. NPV, LR-, and AUC were not significantly different between the 2 filters. These findings informed that an EKG setting in routine practice may be more suitable to predict the presence of myocardial scar because of its higher specificity. Given lower sensitivity, nonetheless, this routine setting should not be used as a screening test.

The presence of f-QRS in the solitary lead revealed a higher sensitivity with a lower specificity, PPV, and LR+ when compared with standard 2 consecutive leads. Although the sensitivity was higher, f-QRS in a single lead was insufficient to be applied as a screening indicator for the presence of myocardial scars.

The mechanism that underlined the formation of f-QRS was an abnormal “zigzag” electrical conduction around the myocardial scars causing multiple spikes (fragmentation, high frequency) on the QRS complex. Myocardial scars are the area of fibrosis containing various types of extracellular substances. Given no viable cardiomyocyte and the presence of dense electrical insulate substances, electrical conduction cannot be conducted through and has to re-route around myocardial scars. (8) However, f-QRS are not recognized only in patients with myocardial scars. One study reported a high prevalence of f-QRS in healthy population (15.6% for inferior leads, 2.9% for anterior leads, and 0.5% for lateral leads) which was not much different from the population with known cardiac disease (16.7% for inferior leads, 3.8% for anterior leads, and 1.8% for lateral leads). (24) The f-QRS was also found to be associated with myocardial ischemia even though absence of scar. (25) In addition, f-QRS was depicted in patients with arrhythmic diseases such as Brugada syndrome and long QT syndrome. The prevalence of f-QRS was as high as 80% when patients presented with ventricular arrhythmia and syncope. (26, 27) These studies suggested that f- QRS were not exclusively specific for myocardial scars.

A few problems regarding the diagnosis of f-QRS are currently unresolved. The present criteria proposed by Das, et al (2006) might include a benign variant of the QRS complex. Some publications have attempted to redefine the definition of pathologic f-QRS; though, no consensus was concluded. (28, 29) Moreover, diagnosing f-QRS was interpreter- dependent and filter-dependent. With the higher low-pass filter, the sensitivity of f-QRS was increased with a lower specificity trade-off. The most appropriate filter in this circumstance was unclear and needs more study to clarify this uncertainty. Most of the previous publications did not state interobserver and intraobserver reliability of f-QRS identification while our study demonstrated interobserver and intraobserver reliability of 0.78 and 0.78 respectively. Despite good agreement indicated by these values, many f-QRS identifications were discordant.

There are several strong points of the current study. This study included the largest number of participants, nearly 1,700 participants, making the results of the study more robust. Moreover, we used 3.0 Tesla CMR as a reference test causing a higher accuracy of myocardial scar diagnosis. All EKGs were recorded just before the CMR scan, minimizing the effect of time on the accuracy of results. Importantly, the current study was the first study that examined the performance of f-QRS between different filter ranges and different numbers of leads.

This study had several limitations that are worth mentioning. Firstly, our study did not investigate the prognostic value of f-QRS. Many studies reported the association between f-QRS and adverse cardiac outcomes in various cardiac diseases. (6, 30–32) Secondly, all EKGs were analyzed on a high-definition display with digital software that can increase the size of EKG waves. With this method of interpretation, the f-QRS detection rate might be more prevalent compared to the paper EKG. This can limit the external generalizability of our result. Thirdly, CMR findings were retrieved from the database and not re-analyzed.

However, all CMR studies in our center were processed and interpreted by cardiac imaging specialists making the reports reliable. Fourthly, the association of f-QRS and the extension and patterns of myocardial scars were not investigated. Knowing these associations may lead to a further understanding of the true value of f-QRS. Lastly, this study did not classify the types of f-QRS. As mentioned before, the exact type of f-QRS that best predicted myocardial scar was unclear.

## CONCLUSION

This study illustrated the limitations of f-QRS and raised the concern about the usage of f-QRS as a marker of myocardial scars. Given its high specificity, however, the presence of f-QRS from EKG performed in routine clinical practice may still be used to rule in the suspected patients for further investigation. We also emphasized the necessity of re-defining the f-QRS diagnostic criteria and validating this criterion with standard scar-detecting modality namely contrast-enhanced CMR.

## Data Availability

Data supporting this study are included within the article and its supplementary materials.

## Acknowledgements

None

---

**Supplementary table 1.** Ischemic vs Non ischemic scars.

|  | Sensitivity<br>(95%CI) | Specificity<br>(95%CI) | PPV<br>(95%CI) | NVP<br>(95%CI) | LR+<br>(95%CI) | LR-<br>(95%CI) | AUC<br>(95%CI) |
| --- | --- | --- | --- | --- | --- | --- | --- |
| Ischemic scars |  |  |  |  |  |  |  |
| Anterior leads and LV segments |  |  |  |  |  |  |  |
| 40-Hz filter | 12.8 (7.5-20) | 96.3 (95-97.3) | 27.6 (16.7-40.9) | 90.9 (89.1-92.5) | 3.45 (2.96-3.94) | 0.91 (0.88-0.94) | 0.55 (0.52-0.58) |
| 100-Hz filter | 20.8 (14.1-29) | 89.1 (87.1-90.8) | 17.3 (11.6-24.4) | 91.1 (89.2-92.7) | 1.9 (1.24-2.56) | 0.89 (0.84-0.94) | 0.55 (0.51-0.59) |
| Lateral leads and LV segments |  |  |  |  |  |  |  |
| 40-Hz filter | 3.1 (0.4-10.7) | 98.2 (97.3-98.9) | 8.7 (1.1-28) | 94.9 (93.5-96.1) | 1.75 (0.97-2.53) | 0.99 (0.97-1.01) | 0.51 (0.49-0.53) |
| 100-Hz filter | 13.8 (6.5-24.7) | 92.5 (90.9-94) | 9.2 (4.3-16.7) | 95.2 (93.8-96.3) | 1.86 (1.21-2.51) | 0.93 (0.9-0.97) | 0.53 (0.49-0.58) |
| Inferior leads and LV segments |  |  |  |  |  |  |  |
| 40-Hz filter | 12.7 (8.9-17.5) | 88.7 (86.5-90.6) | 21.9 (15.5-29.5) | 80.2 (77.8-82.5) | 1.12 (0.94-1.30) | 0.99 (0.94-1.04) | 0.51 (0.48-0.53) |
| 100-Hz filter | 41.7 (35.5-48) | 63.4 (60.4-66.4) | 22.2 (18.5-26.2) | 81.3 (78.4-83.9) | 1.14 (0.99-1.29) | 0.92 (0.89-0.95) | 0.53 (0.49-0.56) |
| Nonischemic scars |  |  |  |  |  |  |  |
| Anterior leads and LV segments |  |  |  |  |  |  |  |
| 40-Hz filter | 14.5 (10.7-19.1) | 96.3 (94.9-97.3) | 51.8 (40.6-62.9) | 80.2 (77.9-82.4) | 3.88 (2.76-4.99) | 0.89 (0.82-0.92) | 0.55 (0.53-0.58) |
| 100-Hz filter | 27 (22.1-32.5) | 88.8 (86.8-90.7) | 40.2 (33.3-47.4) | 81.4 (79.1-83.6) | 2.42 (2.12-2.73) | 0.82 (0.8-0.85) | 0.58 (0.55-0.61) |
| Lateral leads and LV segments |  |  |  |  |  |  |  |
| 40-Hz filter | 8.3 (5-12.7) | 97.6 (96.5-98.4) | 39.1 (25.1-54.6) | 84.8 (82.8-86.7) | 3.38 (2.89-3.87) | 0.94 (0.89-0.97) | 0.53 (0.51-0.55) |
| 100-Hz filter | 24.3 (18.8-30.6) | 92.6 (90.9-94) | 38.4 (30.3-47.1) | 86.5 (84.5-88.4) | 3.27 (2.76-3.78) | 0.82 (0.79-0.86) | 0.58 (0.56-0.61) |
| Inferior leads and LV segments |  |  |  |  |  |  |  |
| 40-Hz filter | 25.5 (20.5-31) | 88.2 (86.1-90) | 36 (29.4-43.1) | 81.9 (79.6-84.1) | 2.16 (1.98-2.34) | 0.85 (0.79-0.91) | 0.57 (0.54-0.6) |
| 100-Hz filter | 43.6 (37.7-49.6) | 63.5 (60.5-66.3) | 23.7 (20.1-27.7) | 81.2 (78.4-83.8) | 1.19 (1.01-1.37) | 0.89 (0.83-0.95) | 0.54 (0.5-0.57) |
| Diagnostic performance of f-QRS for detecting ischemic and non-ischemic scars; LR, likelihood ratio; PPV, positive predictive value; NVP, negative predictive value; OR, odd ratio; CI, confidence interval; Hz, Hertz. |  |  |  |  |  |  |  |

**Supplementary table 2.** Solitary lead.

|  | Sensitivity<br>(95%CI) | Specificity<br>(95%CI) | PPV<br>(95%CI) | NVP<br>(95%CI) | LR+<br>(95%CI) | LR-<br>(95%CI) | AUC<br>(95%CI) |
| --- | --- | --- | --- | --- | --- | --- | --- |
| Anterior leads and LV segments |  |  |  |  |  |  |  |
| 40-Hz filter | 31.8 (27.6-36.4) | 88.7 (86.8-90.4) | 50.5 (44.5-56.5) | 78.3 (76-80.4) | 2.83 (2.61-3.12) | 0.77 (0.69-0.85) | 0.6 (0.58-0.63) |
| 100-Hz filter | 51.2 (46.5-55.9) | 63.1 (60.3-65.8) | 33.4 (29.9-37) | 78.2 (75.5-80.7) | 1.39 (1.22-1.56) | 0.77 (0.67-0.87) | 0.57 (0.55-0.6) |
| Lateral leads and LV segments |  |  |  |  |  |  |  |
| 40-Hz filter | 22.1 (17.6-27.1) | 93 (91.5-94.3) | 41.2 (33.6-49.1) | 84.3 (82.4-86.1) | 3.15 (2.58-3.72) | 0.84 (0.79-0.89) | 0.58 (0.55-0.6) |
| 100-Hz filter | 41.6 (36-47.3) | 76.4 (74-78.6) | 28.1 (24-32.5) | 85.4 (83.4-87.4) | 1.76 (1.27-2.25) | 0.77 (0.70-0.84) | 0.59 (0.56-0.62) |
| Inferior leads and LV segments |  |  |  |  |  |  |  |
| 40-Hz filter | 35.5 (31.5-39.6) | 74.3 (71.6-76.8) | 40.8 (36.4-45.3) | 69.7 (67-72.3) | 1.38 (1.11-1.65) | 0.87 (0.82-0.92) | 0.55 (0.53-0.57) |
| 100-Hz filter | 59.4 (55.2-63.5) | 43.4 (40.4-46.3) | 34.4 (31.4-37.5) | 68.1 (64.6-71.5) | 1.05 (0.94-1.16) | 0.94 (0.89-0.99) | 0.51 (0.49-0.54) |
| Diagnostic performance of f-QRS in the solitary lead for detecting myocardial scars; PPV, positive predictive value; NVP, negative predictive value; OR, odd ratio; CI, confidence interval; Hz, Hertz. |  |  |  |  |  |  |  |

## REFERENCES

1. Longo D, et al. Electrocardiography. Harrison’s principle of internal medicine. 20th ed: McGraw-Hill Education LLC.; 2020. p. 1831-9.

2. Das MK, Zipes DP. Fragmented QRS: a predictor of mortality and sudden cardiac death. Heart Rhythm. 2009;6(3 Suppl):S8-14.

3. Fares H, Heist K, Lavie CJ, Kumbala D, Ventura H, Meadows R, et al. Fragmented QRS complexes-a novel but underutilized electrocardiograhic marker of heart disease. Crit Pathw Cardiol. 2013;12(4):181–3.

4. Das MK, Khan B, Jacob S, Kumar A, Mahenthiran J. Significance of a fragmented QRS complex versus a Q wave in patients with coronary artery disease. Circulation. 2006;113(21):2495–501.

5. Das MK, Saha C, El Masry H, Peng J, Dandamudi G, Mahenthiran J, et al. Fragmented QRS on a 12-lead ECG: a predictor of mortality and cardiac events in patients with coronary artery disease. Heart Rhythm. 2007;4(11):1385–92.

6. Korhonen P, Husa T, Konttila T, Tierala I, Mäkijärvi M, Väänänen H, et al. Fragmented QRS in prediction of cardiac deaths and heart failure hospitalizations after myocardial infarction. Ann Noninvasive Electrocardiol. 2010;15(2):130–7.

7. Sadeghi R, Dabbagh VR, Tayyebi M, Zakavi SR, Ayati N. Diagnostic value of fragmented QRS complex in myocardial scar detection: systematic review and meta-analysis of the literature. Kardiol Pol. 2016;74(4):331–7.

8. Take Y, Morita H. Fragmented QRS: What Is The Meaning? Indian Pacing Electrophysiol J. 2012;12(5):213–25.

9. Das MK, Suradi H, Maskoun W, Michael MA, Shen C, Peng J, et al. Fragmented wide QRS on a 12-lead ECG: a sign of myocardial scar and poor prognosis. Circ Arrhythm Electrophysiol. 2008;1(4):258–68.

10. Mahrholdt H, Wagner A, Judd RM, Sechtem U, Kim RJ. Delayed enhancement cardiovascular magnetic resonance assessment of non-ischaemic cardiomyopathies. Eur Heart J. 2005;26(15):1461–74.

11. Mahenthiran J, Khan BR, Sawada SG, Das MK. Fragmented QRS complexes not typical of a bundle branch block: a marker of greater myocardial perfusion tomography abnormalities in coronary artery disease. J Nucl Cardiol. 2007;14(3):347–53.

12. Dabbagh Kakhki VR, Ayati N, Zakavi SR, Sadeghi R, Tayyebi M, Shariati F. Comparison between fragmented QRS and Q waves in myocardial scar detection using myocardial perfusion single photon emission computed tomography. Kardiol Pol. 2015;73(6):437–44.

13. Kim RJ, Wu E, Rafael A, Chen EL, Parker MA, Simonetti O, et al. The use of contrast-enhanced magnetic resonance imaging to identify reversible myocardial dysfunction. N Engl J Med. 2000;343(20):1445–53.

14. Mann DL. Electrocardiography. Brauwald’s heart disease. 12nd ed. Philadelphia: Elsevier Inc; 2015. p. 114–53.

15. Wagner A, Mahrholdt H, Holly TA, Elliott MD, Regenfus M, Parker M, et al. Contrast- enhanced MRI and routine single photon emission computed tomography (SPECT) perfusion imaging for detection of subendocardial myocardial infarcts: an imaging study. Lancet. 2003;361(9355):374- 9.

16. Cuocolo A, Pace L, Ricciardelli B, Chiariello M, Trimarco B, Salvatore M. Identification of viable myocardium in patients with chronic coronary artery disease: comparison of thallium-201 scintigraphy with reinjection and technetium-99m-methoxyisobutyl isonitrile. J Nucl Med. 1992;33(4):505–11.

17. Marzullo P, Sambuceti G, Parodi O. The role of sestamibi scintigraphy in the radioisotopic assessment of myocardial viability. J Nucl Med. 1992;33(11):1925–30.

18. Crean A, Khan SN, Davies LC, Coulden R, Dutka DP. Assessment of Myocardial Scar; Comparison Between F-FDG PET, CMR and Tc-Sestamibi. Clin Med Cardiol. 2009;3:69–76.

19. Ahn MS, Kim JB, Yoo BS, Lee JW, Lee JH, Youn YJ, et al. Fragmented QRS complexes are not hallmarks of myocardial injury as detected by cardiac magnetic resonance imaging in patients with acute myocardial infarction. Int J Cardiol. 2013;168(3):2008–13.

20. Tian Y, Zhang Y, Yan Q, Mao J, Dong J, Ma C, et al. Fragmented QRS Complex in Healthy Adults: Prevalence, Characteristics, Mechanisms, and Clinical Implications. International Journal of Heart Rhythm. 2017;2(1):34–9.

21. Sarıkaya R, Şengül C, Kümet Ö, İmre G, Akbulut T, Oğuz M. Fragmented QRS in inferior leads is associated with non-alcholic fatty liver disease, body-mass index, and interventricular septum thickness in young men. Anatol J Cardiol. 2022;26(2):100–4.

22. Basaran Y, Tigen K, Karaahmet T, Isiklar I, Cevik C, Gurel E, et al. Fragmented QRS complexes are associated with cardiac fibrosis and significant intraventricular systolic dyssynchrony in nonischemic dilated cardiomyopathy patients with a narrow QRS interval. Echocardiography. 2011;28(1):62–8.

23. Ratheendran AC, Subramanian M, Bhanu DK, Prabhu MA, Kannan R, Natarajan KU, et al. Fragmented QRS on electrocardiography as a predictor of myocardial scar in patients with hypertrophic cardiomyopathy. Acta Cardiol. 2020;75(1):42–6.

24. Terho HK, Tikkanen JT, Junttila JM, Anttonen O, Kenttä TV, Aro AL, et al. Prevalence and prognostic significance of fragmented QRS complex in middle-aged subjects with and without clinical or electrocardiographic evidence of cardiac disease. Am J Cardiol. 2014;114(1):141–7.

25. Hekmat S, Pourafkari L, Ahmadi M, Chavoshi MR, Zamani B, Nader ND. Fragmented QRS on surface electrocardiogram as a predictor of perfusion defect in patients with suspected coronary artery disease undergoing myocardial perfusion imaging. Indian Heart J. 2018;70 Suppl 3(Suppl 3):S177-s81.

26. Morita H, Kusano KF, Miura D, Nagase S, Nakamura K, Morita ST, et al. Fragmented QRS as a marker of conduction abnormality and a predictor of prognosis of Brugada syndrome. Circulation. 2008;118(17):1697–704.

27. Haraoka K, Morita H, Saito Y, Toh N, Miyoshi T, Nishii N, et al. Fragmented QRS is associated with torsades de pointes in patients with acquired long QT syndrome. Heart Rhythm. 2010;7(12):1808–14.

28. Haukilahti MA, Eranti A, Kenttä T, Huikuri HV. QRS Fragmentation Patterns Representing Myocardial Scar Need to Be Separated from Benign Normal Variants: Hypotheses and Proposal for Morphology based Classification. Front Physiol. 2016;7:653.

29. Brohet C. Fragmentation of the QRS complex: the latest electrocardiographic craze? Acta Cardiol. 2019;74(3):185–7.

30. Gong B, Li Z. Total Mortality, Major Adverse Cardiac Events, and Echocardiographic-Derived Cardiac Parameters with Fragmented QRS Complex. Ann Noninvasive Electrocardiol. 2016;21(4):404–12.

31. Güngör B, Özcan KS, Karataş MB, Şahin İ, Öztürk R, Bolca O. Prognostic Value of QRS Fragmentation in Patients with Acute Myocardial Infarction: A Meta-Analysis. Ann Noninvasive Electrocardiol. 2016;21(6):604–12.

32. Jain R, Singh R, Yamini S, Das MK. Fragmented ECG as a risk marker in cardiovascular diseases. Curr Cardiol Rev. 2014;10(3):277–86.

